# Is One Run Enough? Reproducibility of Flagship Large Language Models Across Temperature and Reasoning Settings in Biomedical Text Processing

**DOI:** 10.64898/2026.02.02.26345352

**Authors:** Paul Windisch, Carole Koechli, Fabio Dennstädt, Daniel M. Aebersold, Daniel R. Zwahlen, Robert Förster, Christina Schröder

**Affiliations:** Department of Radiation Oncology, Cantonal Hospital Winterthur, Winterthur, Switzerland; Department of Radiation Oncology, Inselspital, Bern University Hospital, University of Bern, Bern, Switzerland

**Keywords:** Natural language processing, Large language models, Reproducibility, Reasoning models, Temperature

## Abstract

**Purpose:** To quantify run-to-run reproducibility of Gemini 3 Flash Preview and GPT-5.2 for biomedical trial-success classification across temperature and reasoning/thinking settings, and to assess whether single-run reporting is sufficient.

**Methods:** We utilized 250 randomized controlled oncology trial abstracts labeled POSITIVE/NEGATIVE based on primary endpoint success. With a fixed prompt requiring exactly “POSITIVE” or “NEGATIVE”, we evaluated Gemini across thinking levels (minimal, low, medium, high) and temperatures 0.0 - 2.0, and GPT-5.2 across reasoning-effort levels (none to xhigh) with an additional temperature sweep when reasoning was disabled. Each setting was run three times. Reproducibility was quantified with Fleiss’ κ across replicates, performance was summarized with F1 (per run and majority vote), and invalid-format outputs were recorded.

**Results:** Gemini showed near-perfect agreement across settings (κ=0.942 - 1.000), including perfect agreement at temperature 0. Invalid outputs were uncommon (0 - 1.5%). GPT-5.2 reproducibility was similarly high (κ=0.984 - 0.995) with no invalid outputs. Performance remained stable (mean/majority-vote F1 = 0.955 - 0.971), and majority voting offered only marginal gains.

**Conclusion:** For strict binary biomedical classification with tightly constrained outputs, both models were highly reproducible across common decoding and reasoning configurations, indicating that one run is often adequate while minimal replication provides a practical stability check.

## Introduction

Large language models (LLMs) are increasingly used for biomedical text classification and information extraction, including clinical note phenotyping, trial eligibility screening, and systematic review support.^1–3^ Reported performance in these studies is typically summarized using metrics such as accuracy or F1 score, often without explicit consideration of run-to-run variability or reproducibility under identical experimental conditions.^4^ This practice contrasts with established norms in biomedical research, where robustness and repeatability are essential for methodological credibility and downstream clinical translation.^5^

Recent biomedical evaluations of LLMs have demonstrated strong average performance across a range of natural language processing tasks, including structured data extraction and document classification.^6^ However, most studies either implicitly assume deterministic behavior or report results from a single model invocation per input, despite the fact that modern LLMs employ probabilistic decoding and, increasingly, adaptive reasoning mechanisms (e.g., temperature-based sampling or variable “reasoning effort”) that may introduce stochastic variability into outputs.^7^

Several recent works have begun to acknowledge this issue by reporting repeated evaluations or inter-run agreement metrics.^8,9^ For example, reproducibility across repeated LLM invocations has been assessed for clinical text extraction using agreement statistics such as Cohen’s κ or Krippendorff’s α, with findings suggesting that reproducibility may decrease as decoding stochasticity increases.^8,10^ Similarly, studies in guideline adherence and clinical reasoning have shown that repeated queries to the same model can yield divergent outputs, with agreement ranging from near-perfect to effectively random depending on task formulation and prompting strategy.^11,12^ While these studies highlight the existence of variability, they do not provide systematic guidance on whether multiple runs are necessary for reliable reporting, nor do they jointly evaluate the effects of sampling stochasticity and reasoning configuration.

At the same time, LLM APIs have evolved to expose explicit controls over both sampling behavior (e.g., temperature) and inference-time reasoning behavior (e.g., “reasoning effort” or “thinking level”). These controls are increasingly used in biomedical research workflows but remain poorly characterized from a reproducibility standpoint. Importantly, existing biomedical studies typically examine either temperature effects or reasoning mechanisms in isolation, focus on performance rather than reproducibility as a primary endpoint, or restrict evaluation to a single parameter setting per model. To date, there has been no systematic biomedical evaluation that quantifies run-to-run reproducibility across both temperature and reasoning-effort settings for contemporary flagship LLMs.

This gap has practical implications for biomedical publishing. Reviewers increasingly ask whether reported LLM performance metrics are based on a single run or multiple runs, yet there is no empirical consensus on whether repeated runs materially change conclusions under commonly used settings. If single-run evaluations are sufficient under certain configurations, unnecessary replication inflates computational cost and environmental burden without scientific benefit.^13^ Conversely, if multiple runs are required to achieve stable estimates, single-run reporting risks overstating performance and undermining reproducibility.^4,13^

In this study, we therefore conduct a systematic evaluation of LLM reproducibility as a function of decoding temperature and reasoning configuration. Using Gemini Flash with reasoning levels set to minimal, low, medium, and high across a temperature range of 0 to 2.0, we assess run-to-run reproducibility over three repeated invocations per setting. As a comparator, we evaluate OpenAI’s GPT-5.2 across all supported reasoning settings, with additional temperature sweeps performed when reasoning is disabled. Reproducibility is treated as the primary endpoint, while task performance (F1 score) is reported as a secondary metric to contextualize reproducibility-performance trade-offs.

By explicitly addressing the question of whether single-run evaluation is sufficient under modern LLM configurations, this work aims to provide empirically grounded guidance for biomedical researchers and reviewers. Our findings are intended to inform best practices for reporting LLM-based results, balancing scientific rigor against computational efficiency, and to reduce the risk of publishing irreproducible findings in biomedical natural language processing.

## Methods

This study evaluated run-to-run reproducibility of LLMs for a biomedical text classification task using an existing, previously published dataset from the same author group. The dataset consists of 250 randomized controlled oncology trials (RCTs) drawn from seven major medical journals (British Medical Journal, JAMA, JAMA Oncology, Journal of Clinical Oncology, Lancet, Lancet Oncology, and New England Journal of Medicine) published between 2005 and 2023. In the original publication, eligible trials were restricted to designs with exactly two arms and a single primary endpoint, and abstracts were retrieved via PubMed and parsed from text to create the study corpus. The rationale for these restrictions was to reduce edge cases and ensure a consistent definition of “trial success”. For the present reproducibility study, we reused the released dataset and corresponding ground-truth labels in local CSV format.

Ground-truth labels were adopted from the original dual-annotation procedure performed by two authors (P.W., C.K.), in which trials were classified as positive if the primary endpoint was met and negative otherwise. The annotation workflow in the original study included an initial calibration phase, followed by independent labeling and consensus resolution of discrepancies. A third reviewer was designated for unresolved conflicts but was not needed (D.R.Z.). Annotation was conducted using Prodigy, and full texts or protocols were consulted only when the abstract did not clearly report the primary endpoint and its results. We did not modify labels or repeat manual annotation for the present analysis.

The task for the LLMs was to determine, from the provided trial text, whether the trial met its primary endpoint. To align with the previously published workflow and minimize ambiguity, we used an explicit instruction format adapted from the original study’s prompt design, requiring the model to output exactly one token-level label: POSITIVE or NEGATIVE in all caps. In contrast to the earlier feasibility study—which intentionally refrained from repeated classification calls under its chosen settings—here we explicitly treated reproducibility as a primary endpoint and therefore repeated inference multiple times under identical conditions. Two commercial LLMs were evaluated via their vendor APIs in a local pipeline: OpenAI’s GPT-5.2 (snapshot gpt-5.2-2025-12-11) and Google’s Gemini 3 Flash (model gemini-3-flash-preview). In accordance with the study design, we evaluated decoding temperature and model “reasoning/thinking” controls across all supported levels. Gemini 3 Flash was evaluated across all available thinking levels (minimal, low, medium, high) and a temperature grid of 0, 0.5, 1.0, 1.5, and 2.0, using every permutation. GPT-5.2 was evaluated across all supported reasoning-effort levels (none, low, medium, high, xhigh). Because GPT-5.2 only permits user-specified temperature when reasoning effort is set to none, the full temperature sweep was performed only under reasoning=none, while the remaining reasoning settings were evaluated without specifying temperature. This constraint reflects current API behavior. No other decoding parameters were set and no seed was specified, in order to capture natural run-to-run variability.

We ran the whole dataset three times, yielding three replicate labels per trial/setting combination.

Responses were considered valid only if the returned text matched exactly POSITIVE or NEGATIVE after trimming whitespace. Any other output was recorded as invalid for downstream summaries.

The following system prompt (i.e., the fixed instruction provided to the model to define its task) was used: “You will be provided with the title and abstract of a randomized controlled oncology trial. Your task will be to classify if the trial was positive, i.e. if it met its primary endpoint, or negative, i.e. if it did not meet its primary endpoint. Your response should be either the word POSITIVE (in all caps) or NEGATIVE (in all caps).”

The user prompt (i.e., the specific input text) was the respective title and abstract

### Statistical analysis

Reproducibility was the main endpoint and was quantified per setting using Fleiss’ κ computed across the three replicate outputs over all trials. Task performance was evaluated as a secondary endpoint to contextualize reproducibility-performance trade-offs. For the analysis of performance we calculated for each combination of model and setting the F1 score per run, the mean F1 score for after all runs, and the F1 score based on a majority vote for each trial based on all runs. For the majority vote, if at least two of three replicates agreed on a valid label, that label was used as the final prediction. For the calculation of the F1 score, only valid, i.e. correctly formatted predictions were considered.

### Ethical consideration

This study used publicly available abstracts from published clinical trials and contained no patient-level information. Therefore, ethics approval was not required.

## Results

A total of 250 oncology RCT abstracts were evaluated, with three replicate model invocations per setting.

For Gemini 3 Flash Preview, run-to-run reproducibility was consistently high across thinking levels and temperatures (Fleiss’ κ, 0.942 - 1.000, Table 1). Perfect agreement was observed at temperature 0.0 across all thinking levels (κ=1.000). The lowest agreement occurred at medium thinking and temperature 2.0 (κ=0.942). Performance was stable across settings, with mean F1 scores ranging from 0.955 to 0.966 and majority-vote F1 scores ranging from 0.955 to 0.967. Within-setting F1 ranges across the three runs were narrow (maximum spread 0.012). Invalid outputs were uncommon (0-1.5%).

**Table 1.** Reproducibility and Performance of Gemini 3 Flash Preview at different reasoning efforts and temperature settings.

| Reasoning level | Temperature | Fleiss' Kappa | F1 mean | F1 range | F1 majority | Invalid [%] |
| --- | --- | --- | --- | --- | --- | --- |
| Minimal | 0.0 | 1.0 | 0.963 | 0.963 - 0.963 | 0.963 | 0.0 |
|  | 0.5 | 0.995 | 0.963 | 0.963 - 0.963 | 0.963 | 0.1 |
|  | 1.0 | 0.979 | 0.963 | 0.963 - 0.963 | 0.963 | 0.8 |
|  | 1.5 | 0.968 | 0.963 | 0.963 - 0.963 | 0.963 | 1.2 |
|  | 2.0 | 0.979 | 0.963 | 0.963 - 0.963 | 0.963 | 0.8 |
| High | 0.0 | 1.0 | 0.955 | 0.955 - 0.955 | 0.955 | 0.0 |
|  | 0.5 | 0.989 | 0.958 | 0.955 - 0.959 | 0.959 | 0.3 |
|  | 1.0 | 0.995 | 0.956 | 0.955 - 0.959 | 0.955 | 0.4 |
|  | 1.5 | 0.984 | 0.959 | 0.955 - 0.963 | 0.959 | 0.4 |
|  | 2.0 | 0.97 | 0.96 | 0.955 - 0.967 | 0.959 | 0.5 |
| Medium | 0.0 | 1.0 | 0.959 | 0.959 - 0.959 | 0.959 | 0.8 |
|  | 0.5 | 0.973 | 0.966 | 0.963 - 0.967 | 0.963 | 0.7 |
|  | 1.0 | 0.963 | 0.961 | 0.959 - 0.967 | 0.959 | 0.8 |
|  | 1.5 | 0.952 | 0.965 | 0.963 - 0.967 | 0.963 | 1.2 |
|  | 2.0 | 0.942 | 0.964 | 0.963 - 0.967 | 0.963 | 1.5 |
| High | 0.0 | 1.0 | 0.963 | 0.963 - 0.963 | 0.963 | 0.8 |
|  | 0.5 | 0.952 | 0.964 | 0.962 - 0.967 | 0.967 | 0.9 |
|  | 1.0 | 0.962 | 0.96 | 0.959 - 0.963 | 0.959 | 0.7 |
|  | 1.5 | 0.952 | 0.961 | 0.958 - 0.963 | 0.963 | 1.3 |
|  | 2.0 | 0.962 | 0.964 | 0.963 - 0.967 | 0.967 | 0.7 |

For GPT-5.2, reproducibility remained high across all evaluated reasoning-effort settings (Fleiss’ κ, 0.984 - 0.995, Table 2). With reasoning disabled (none), κ ranged from 0.984 to 0.995 across temperatures 0.0-2.0, with majority-vote F1 scores of 0.963 - 0.967. Across reasoning-effort levels low through xhigh (temperature not user-specified), κ ranged from 0.984 to 0.995 and majority-vote F1 from 0.959 to 0.971. No invalid outputs were observed.

**Table 2.** Reproducibility and Performance of GPT-5.2 at different reasoning efforts and temperature settings.

| Reasoning level | Temperature | Fleiss' Kappa | F1 mean | F1 range | F1 majority | Invalid [%] |
| --- | --- | --- | --- | --- | --- | --- |
| **None** | 0.0 | 0.984 | 0.964 | 0.959 - 0.971 | 0.963 | 0.0 |
|  | 0.5 | 0.989 | 0.963 | 0.963 - 0.963 | 0.963 | 0.0 |
|  | 1.0 | 0.995 | 0.966 | 0.963 - 0.967 | 0.967 | 0.0 |
|  | 1.5 | 0.989 | 0.963 | 0.963 - 0.963 | 0.963 | 0.0 |
|  | 2.0 | 0.995 | 0.966 | 0.963 - 0.967 | 0.967 | 0.0 |
| **Low** |  | 0.984 | 0.963 | 0.963 - 0.963 | 0.959 | 0.0 |
| **Medium** |  | 0.995 | 0.964 | 0.963 - 0.967 | 0.963 | 0.0 |
| **High** |  | 0.995 | 0.969 | 0.967 - 0.971 | 0.967 | 0.0 |
| **Xhigh** |  | 0.995 | 0.97 | 0.967 - 0.971 | 0.971 | 0.0 |

For GPT-5.2, the mean F1 scores increased with increased reasoning effort, while this was not the case for Gemini 3 Flash Preview (Figure 1).

**Figure 1.**
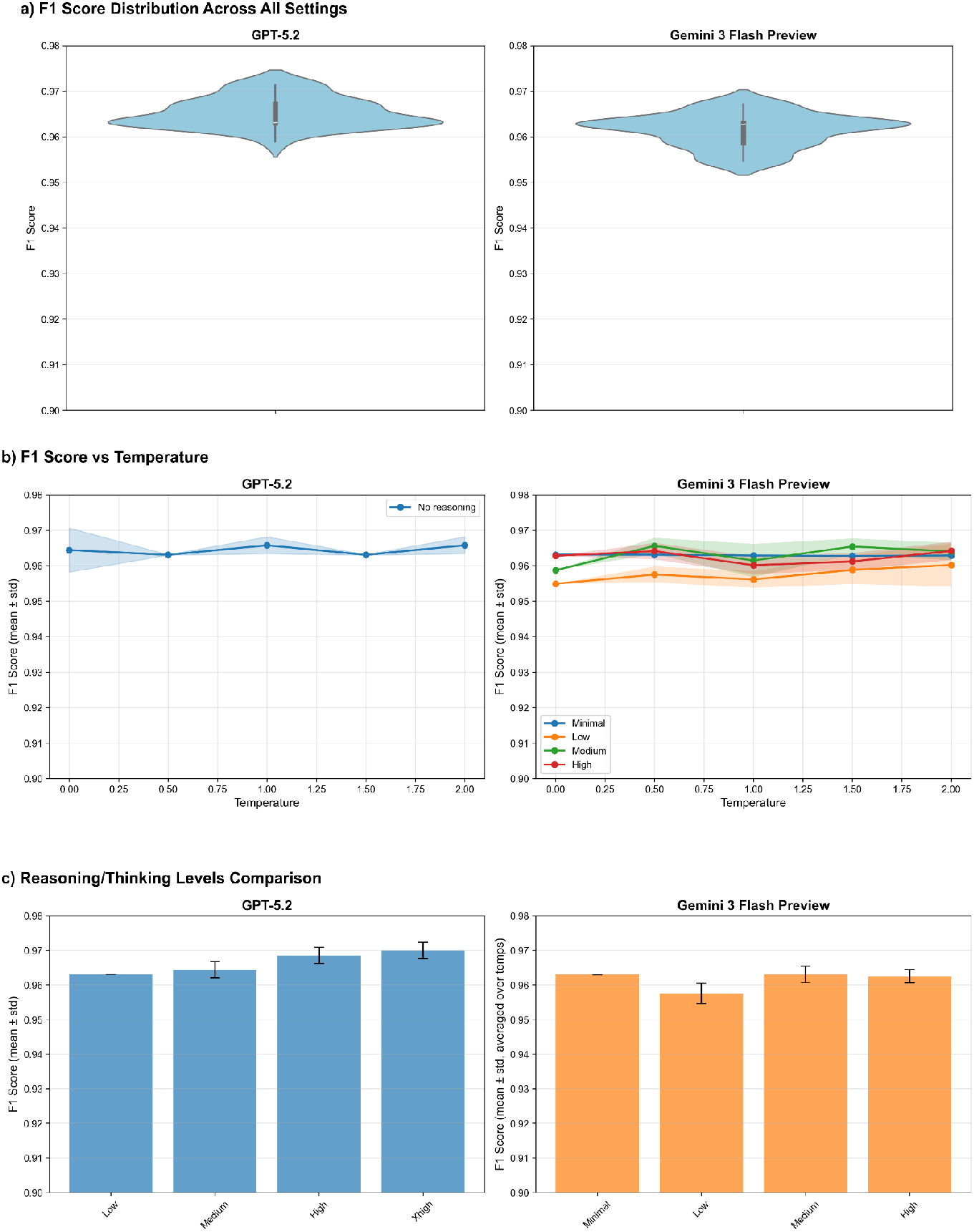
Combined performance for GPT-5.2 (left) and Gemini 3 Flash Preview right. a) F1 scores across all settings. b) F1 score by temperature per reasoning setting. Translucent bands indicate the F1 score ranges. c) F1-scores by reasoning effort. Error bars indicate the F1 score ranges.

Analysis of the confusion matrices for the setting combination with the greatest performance differences between runs for both GPT-5.2 and Gemini 3 Flash Preview did not reveal a clear pattern of failure Figure 2).

**Figure 2.**
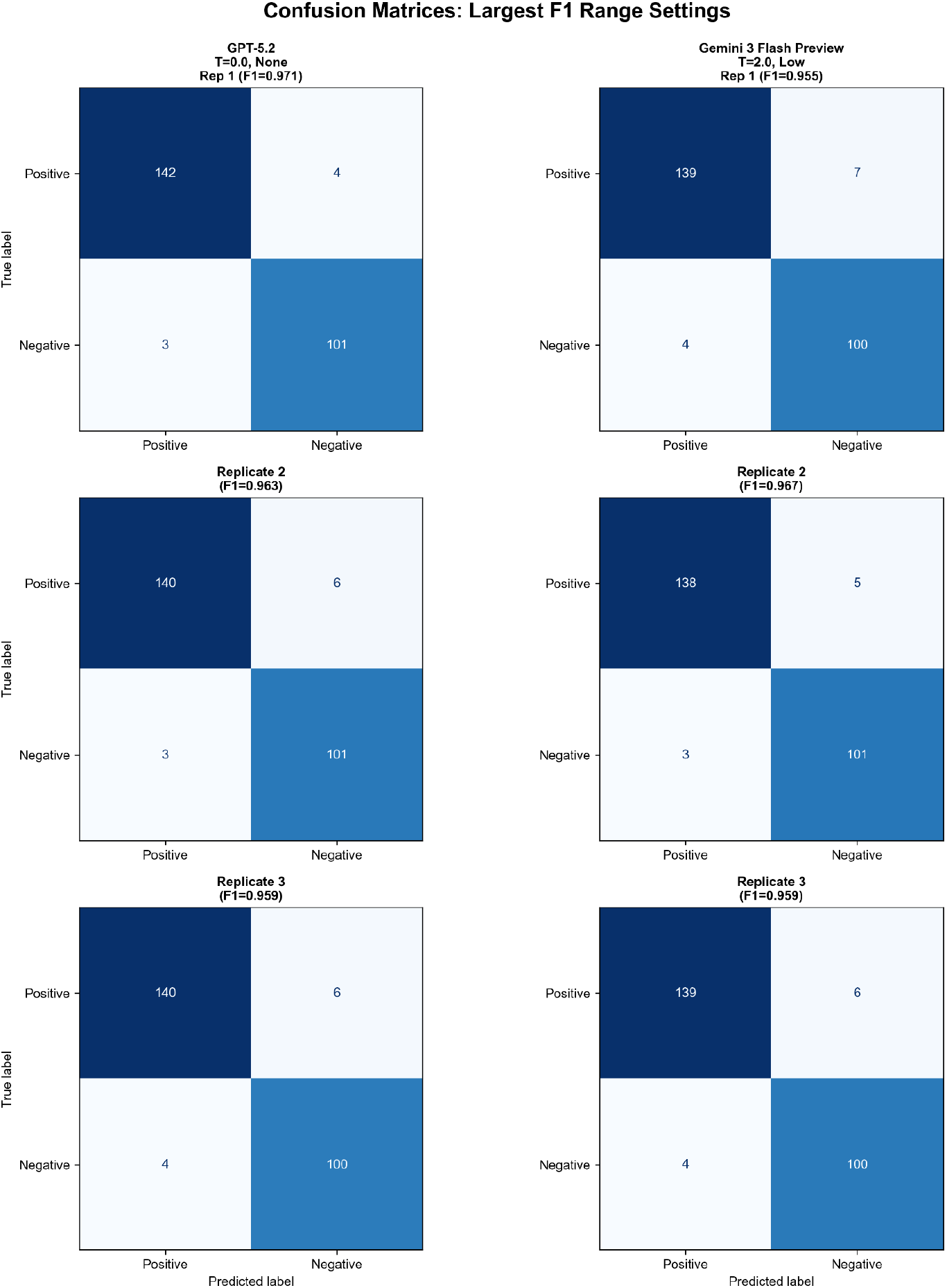
Confusion matrices for model and setting combination with the greatest difference between best and worst F1 score for GPT-5.2 (left) and Gemini 3 Flash Preview (right).

## Discussion

Across 250 oncology RCT abstracts, both Gemini 3 Flash Preview and GPT-5.2 showed consistently high run-to-run reproducibility for binary “trial success” classification when the input and prompts were held constant. For Gemini 3 Flash Preview, agreement was near-perfect across all thinking levels and temperatures (Fleiss’ κ 0.942 - 1.000), with perfect agreement at temperature 0.0 and only modest decreases at the highest temperature settings. For GPT-5.2, reproducibility likewise remained high across all reasoning-effort settings and across the temperature sweep when reasoning was disabled (Fleiss’ κ 0.984 - 0.995). Task performance (F1) was stable across runs within each setting, and majority voting over three runs yielded only small improvements relative to single-run results, indicating that—at least for this constrained binary endpoint—repeating runs primarily adds reassurance and a quantifiable stability estimate rather than materially changing conclusions.

These findings add nuance to a growing literature warning that single-run evaluations of generative AI can be misleading because outputs may vary stochastically even under “identical” conditions. Zhu and colleagues explicitly argue that evaluation should include repetition to assess stability.^4^ In clinical settings, run-to-run variability has been demonstrated most clearly in open-ended or recommendation-style tasks, where multiple reasonable continuations exist and where prompting can elicit different management choices. For example, Landon et al. reported substantial variation in LLM recommendations for challenging inpatient management scenarios and explicitly advised clinicians to treat any single output as only one perspective.^11^ Similarly, guideline-concordance work has highlighted internal consistency problems and low agreement with reference recommendations in some domains, underscoring that “accuracy” alone may not capture reliability.^12^ In contrast, our task is deliberately constrained (exactly one of two labels), and the evaluation prompt enforces a narrow output space—conditions that likely reduce expressive degrees of freedom and therefore suppress variability. This interpretation is consistent with prior biomedical extraction/classification evaluations in which repeated same-prompt iterations produced very high agreement metrics (e.g., Krippendorff’s α close to 1 across several high-performing models in a multi-task EHR extraction study).^8^ It is also consistent with repeated-query studies in medical education contexts, which typically observe non-trivial but task-dependent consistency rather than guaranteed determinism, even when questions are objective.^14,15^

With respect to temperature and reasoning controls, our results align with the conceptual expectation that higher temperature can increase sampling diversity and potentially reduce stability.^7^ We did observe the lowest agreement for Gemini at the most stochastic settings, but the magnitude of the drop remained small in absolute terms for this binary task. The practical implication is that “temperature-induced instability” may be strongly task- and output-format dependent: where the correct decision boundary is clear from the input (as is often the case for trial abstracts explicitly reporting whether the primary endpoint was met), different sampled continuations may still collapse onto the same final label. Notably, GPT-5.2 showed improved mean F1 with increased reasoning effort without an accompanying loss of reproducibility. This suggests that, in at least some contemporary models, allocating more inference-time reasoning may improve decision quality while preserving run-to-run stability. However, given that the GPT-5.2 API constraint prevented an explicit temperature sweep when reasoning was enabled, disentangling the independent contributions of reasoning effort versus implicit/default decoding behavior remains an important consideration for interpretation and for future benchmarking.

Several features strengthen the interpretability and practical value of this work. First, we treated reproducibility as a primary outcome rather than an afterthought, using a standard chance-corrected agreement statistic (Fleiss’ κ) over repeated invocations and reporting invalid-output rates alongside performance. Second, we evaluated two widely used commercial model families and systematically swept temperature and reasoning/thinking levels across the full range supported by each vendor, producing a configuration map that is directly actionable for researchers designing biomedical NLP pipelines. Third, the dataset covers 250 oncology RCTs sampled across multiple high-impact journals and nearly two decades of publication, which increases topical and stylistic diversity relative to single-source corpora. Finally, strict output validation (POSITIVE/NEGATIVE only) makes failure modes transparent and helps separate “format reliability” from “classification correctness,” an operational distinction that matters for downstream automation.

At the same time, several limitations constrain generalizability. The study evaluates one binary classification task with a tightly restricted output space, so results should not be assumed to extend to open-ended generation (e.g., clinical recommendations, narrative summarization, or free-form data extraction), where multiple plausible outputs can be semantically different yet superficially acceptable. Evidence from other domains suggests that variability can be larger in precisely those less constrained setting.^11,16^ Reproducibility was assessed within a single model snapshot and execution period. It does not address longitudinal reproducibility across model updates, infrastructure changes, or silent backend modifications, which are increasingly recognized as practical threats to repeatability. Recent work in adjacent educational evaluation settings underscores that stability and performance can change across sessions and over time, and that “single-trial accuracy can mask volatility and stable systematic errors”.^17^ In addition, GPT-5.2’s inability to specify temperature when reasoning is enabled means that comparisons between reasoning levels are potentially confounded by unobserved defaults. Finally, the dataset itself was curated to reduce ambiguity (two-arm RCTs, single primary endpoint), which likely increases determinism relative to messier real-world inputs (e.g., heterogeneous trial designs, multi-endpoint reporting, or incomplete abstracts). This choice was appropriate for isolating run-to-run effects, but it may overestimate reproducibility relative to broader biomedical text mining deployments.

Future work should test whether the “one run is enough” conclusion holds across (i) more linguistically and clinically heterogeneous tasks (entity extraction, multi-label classification, evidence summarization), (ii) less constrained output formats (structured JSON with multiple fields, rationale + label, or chain-of-thought-free but explanation-bearing outputs), and (iii) more challenging inputs where ambiguity is intrinsic rather than curated away. Methodologically, studies could evaluate adaptive replication policies (e.g., run once by default, but trigger additional runs when model confidence is low or when a first-pass ensemble disagrees) to balance rigor with compute and environmental cost.

In conclusion, for a constrained binary biomedical text classification problem with strict output formatting, both evaluated flagship LLMs exhibited near-perfect run-to-run reproducibility across a broad range of temperature and reasoning/thinking settings, and majority voting over three runs offered minimal performance gains. This suggests that single-run reporting may often be adequate for similar tasks and prompts, particularly at low-to-moderate stochasticity, while not challenging the broader recommendation that researchers should explicitly report decoding and reasoning configurations and—where feasible—include at least a minimal replication-based stability check to guard against hidden variability and to enable trustworthy comparison across studies.

## Ethics approval and consent to participate

Not applicable

## Availability of data and materials

All data and code used to obtain this study’s results have been uploaded to https://github.com/windisch-paul/llm_reproducibility

## Author contributions

Conceptualization, P.W., F.D.; methodology, P.W, C.S.; formal analysis, P.W.; data curation, P.W., C.K.; writing—original draft preparation, P.W., C.S.; writing—review and editing, C.K., F.D., D.M.A., D.R.Z., R.F.; supervision, D.R.Z.; project administration, D.R.Z.;

All authors read and approved the final manuscript.

